# Diabetes as a Driver of Financial Toxicity Among U.S. Cancer Survivors: A Nationally Representative Analysis of NHIS 2021–2024

**DOI:** 10.64898/2026.07.12.26357889

**Authors:** Jay Tewari, Vanshika Singh, Khalid Ahmad Qidwai, Abhijay Shah, Ajoy Tewari, Vineeta Tewari, Harshit Narula

**Affiliations:** Internal Medicine PGY-1, Baptist Hospitals of Southeast Texas, Beaumont, Texas, USA; Internal Medicine, King George’s Medical University, Lucknow, India; Internal Medicine PGY-1, Cleveland Clinic Akron General Hospital, Akron, Ohio, USA; Internal Medicine, Hind Institute of Medical Sciences, Barabanki, India; Anatomy, Era’s Lucknow Medical College and Hospital, Lucknow, India; Internal Medicine, Sinai Hospital of Baltimore, Baltimore, Maryland, USA

## Abstract

**Background:** Financial toxicity is an increasingly recognized survivorship issue, but whether diabetes identifies a distinct high-risk financial-toxicity phenotype among U.S. cancer survivors is not well characterized.

**Methods:** We conducted a cross-sectional study using pooled 2021–2024 National Health Interview Survey Sample Adult data. Adults were classified into four mutually exclusive groups: neither cancer nor diabetes, diabetes only, cancer only, and cancer plus diabetes. The primary outcome was any financial toxicity, defined as cost-related care disruption or medication underuse in the prior 12 months. Survey-weighted prevalence estimates and multivariable Poisson regression were used to calculate adjusted prevalence ratios (aPRs).

**Results:** The weighted analytic population included 210.4 million adults with neither condition, 20.6 million with diabetes only, 20.9 million with cancer only, and 4.3 million with both cancer and diabetes. Any financial toxicity was present in 18.8%, 18.8%, 11.6%, and 17.2% of these groups, respectively. Among cancer survivors, diabetes was associated with higher prevalence of any financial toxicity (aPR 1.51, 95% CI 1.33–1.73), inability to afford prescriptions (aPR 1.71, 95% CI 1.40–2.08), skipped medication doses (aPR 1.91, 95% CI 1.48–2.46), any emergency department visit (aPR 1.35, 95% CI 1.24–1.47), and ≥2 emergency department visits (aPR 1.55, 95% CI 1.32–1.83). In treatment-stratified analyses, the burden was greatest among insulin-treated survivors.

**Conclusions:** Cancer survivors with diabetes represent a high-risk financial-toxicity phenotype despite frequent healthcare contact.

**Impact:** These findings identify a vulnerable survivorship subgroup and support targeted financial navigation, medication affordability interventions, and integrated survivorship-diabetes care strategies.

## Introduction

Financial toxicity is now recognized as an important consequence of cancer survivorship and includes the material, behavioral, and psychological burden created by the cost of care. Cancer survivors are more likely than individuals without cancer to experience debt, distress, delayed care, and treatment nonadherence related to cost (1,2).

Diabetes is especially relevant in this context. It is associated with high direct medical spending, chronic medication use, and ongoing healthcare needs in the United States (3).

Cost-related medication nonadherence is also common in diabetes and may worsen long-term disease control and outcomes (4).

Cancer and diabetes frequently coexist, and the combination can complicate treatment, self-management, and healthcare utilization. Patients living with both conditions report difficulty coordinating care and maintaining diabetes self-management during cancer treatment (5,6). Systematic reviews have shown that people with both cancer and diabetes have worse patient-reported outcomes, and cancer survivors with chronic comorbidities experience greater economic burden than survivors without such conditions (7–9). More recent studies suggest that concurrent diabetes is associated with greater healthcare use and higher economic burden among adults with cancer (10,11).

However, nationally representative data examining whether diabetes identifies a distinct financial-toxicity phenotype among cancer survivors remain limited. We therefore used pooled 2021–2024 National Health Interview Survey (NHIS) data to evaluate financial toxicity and healthcare vulnerability among U.S. adults with neither condition, diabetes only, cancer only, and both cancer and diabetes. We hypothesized that cancer survivors with diabetes would experience greater financial toxicity than cancer survivors without diabetes despite frequent healthcare contact.

## Materials and Methods

### Study design and data source

We conducted a cross-sectional study using pooled 2021–2024 NHIS Sample Adult data. NHIS is a nationally representative, multistage probability survey of the noninstitutionalized U.S. population conducted by the National Center for Health Statistics. The Sample Adult files include annual core measures on chronic conditions, health status, access to care, and healthcare utilization, and the post-2019 redesign maintained broadly comparable adult core content across recent years (12). Because NHIS uses a complex survey design, analyses must account for survey weights, strata, and primary sampling units (12,13).

### Study population

Adults aged 18 years or older in the 2021–2024 NHIS Sample Adult files were eligible. Participants were classified into four mutually exclusive phenotype groups based on self-reported history of cancer and diabetes: neither cancer nor diabetes, diabetes only, cancer only, and cancer plus diabetes. Cancer history was defined using the adult cancer item indicating whether a respondent had ever been told by a health professional that they had cancer or a malignancy of any kind (14). Diabetes was defined using the integrated IPUMS NHIS diabetes-history variable. Respondents with borderline diabetes were excluded from the primary diabetes definition.

### Outcomes

The primary outcome was medical financial toxicity (FT), defined using six NHIS cost-related items assessed over the prior 12 months: delayed medical care due to cost, inability to afford needed medical care, inability to afford prescription medication, delayed filling a prescription to save money, taking less medication to save money, and skipping medication doses to save money. Each item was coded as binary (yes/no). We then derived three related metrics: any FT, defined as at least one affirmative response; FT count, defined as the total number of affirmative items (range 0–6); and FT severity, categorized as none (0 items), low/moderate (1 item), or high (≥2 items). The threshold of ≥2 items was used to identify respondents experiencing multiple concurrent cost-related barriers. Variable definitions are provided in Supplementary Table S1.

Secondary outcomes included healthcare access and utilization measures: no usual place for medical care, any emergency department visit in the prior 12 months, frequent emergency department use (≥2 visits), wellness visit within the prior 12 months, and whether the most recent doctor visit was a wellness visit. “No usual place for care” was defined as reporting no usual source of medical care.

### Covariates

We adjusted for age, sex, race/ethnicity, education, poverty category, insurance category, U.S. region, BMI category, smoking status, self-rated health, cardiovascular risk burden, food security status, and urban-rural classification. Cardiovascular risk burden was defined using self-reported hypertension, high cholesterol, angina, coronary heart disease, myocardial infarction, and stroke.

### Statistical analysis

Survey weights were pooled across four years by dividing the Sample Adult weight by four, consistent with NHIS and IPUMS guidance for pooled analyses using R (12,13). Weighted descriptive estimates were used to calculate nationally representative population counts and prevalences. Survey-weighted Poisson regression with robust variance estimation was used to calculate adjusted prevalence ratios (aPRs) and 95% confidence intervals. Primary analyses compared the four phenotype groups, using adults with neither cancer nor diabetes as the reference group. Secondary analyses were restricted to cancer survivors and compared those with versus without diabetes. Prespecified subgroup analyses among cancer survivors evaluated heterogeneity by age group, sex, insurance status, and poverty category. To examine whether treatment intensity identified a gradient of risk, cancer survivors with diabetes were further stratified into those with no current diabetes medications reported, non-insulin-treated diabetes, and insulin-treated diabetes. Additional analyses were performed to address social and geographic vulnerability. We examined food insecurity across the four phenotype groups, reran the primary FT severity model with additional adjustment for food security and urban-rural classification, and performed an urban-rural stratified sensitivity analysis. Exploratory site-specific analyses were also performed for selected common cancer types. Adjusted models were estimated using complete-case analysis; respondents with missing data on any variable included in a given model were excluded from that model. Variable-specific missingness and complete-case sample sizes are summarized in Supplementary Table S6. Detailed supplementary analyses are shown in Supplementary Tables S2–S5.

## Results

### Study population

The weighted analytic population included four mutually exclusive groups: 210.4 million adults with neither cancer nor diabetes, 20.6 million with diabetes only, 20.9 million with cancer only, and 4.3 million with both cancer and diabetes. The cancer-plus-diabetes group was therefore the smallest but clinically distinct subgroup.

In descriptive analyses, mean ages were 44.8 years in the neither group, 60.4 years in the diabetes-only group, 65.2 years in the cancer-only group, and 69.2 years in the cancer-plus-diabetes group. The cancer-only group was predominantly non-Hispanic White, whereas the diabetes-only and cancer-plus-diabetes groups showed greater sociodemographic vulnerability, including lower educational attainment, greater poverty, poorer self-rated health, higher cardiometabolic burden, and greater food insecurity.

### Crude prevalence of financial toxicity across phenotype groups

The prevalence of any financial toxicity was 18.8% among adults with neither condition, 18.8% among those with diabetes only, 11.6% among those with cancer only, and 17.2% among those with both cancer and diabetes (Table 2; Figure 1). Although crude FT prevalence was lower in the cancer-only group than in the neither group, the cancer-only group was substantially older and more likely to have Medicare coverage. After multivariable adjustment, this apparent crude difference attenuated, whereas diabetes remained strongly associated with greater FT burden.

**Table 1.**
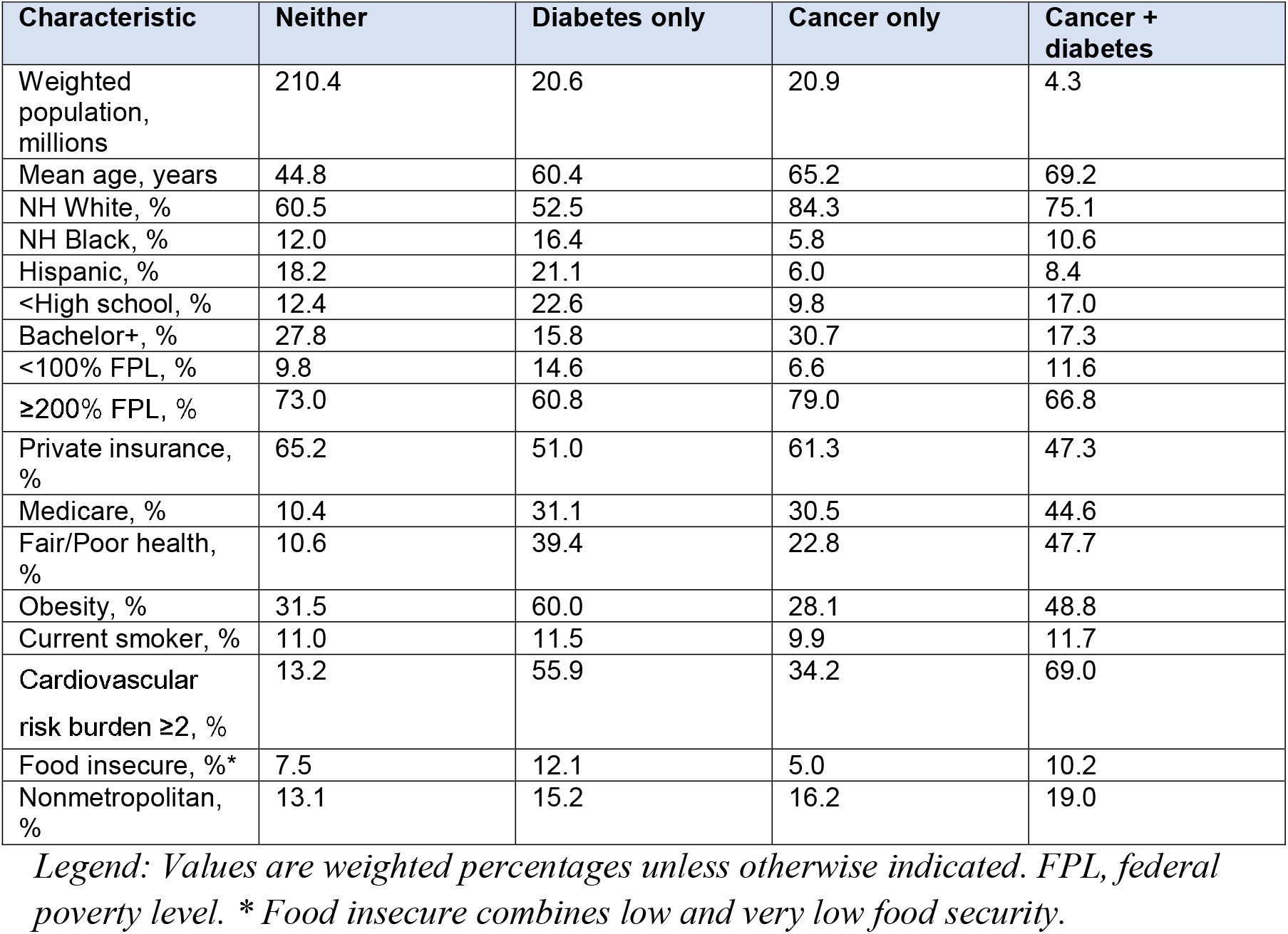
Selected baseline characteristics by cancer-diabetes phenotype.

**Table 2.**
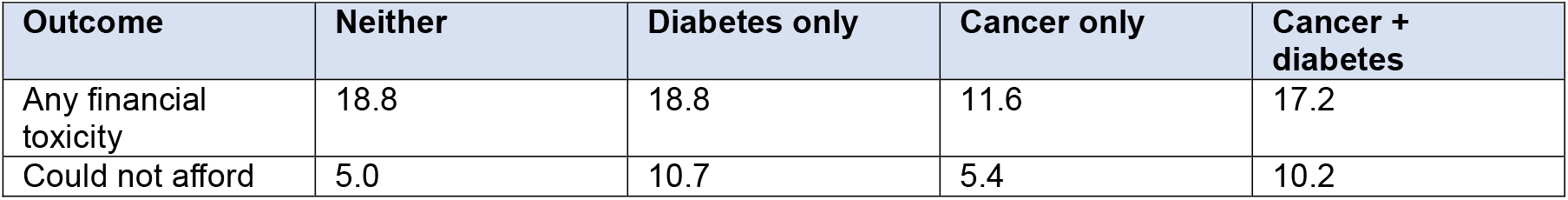
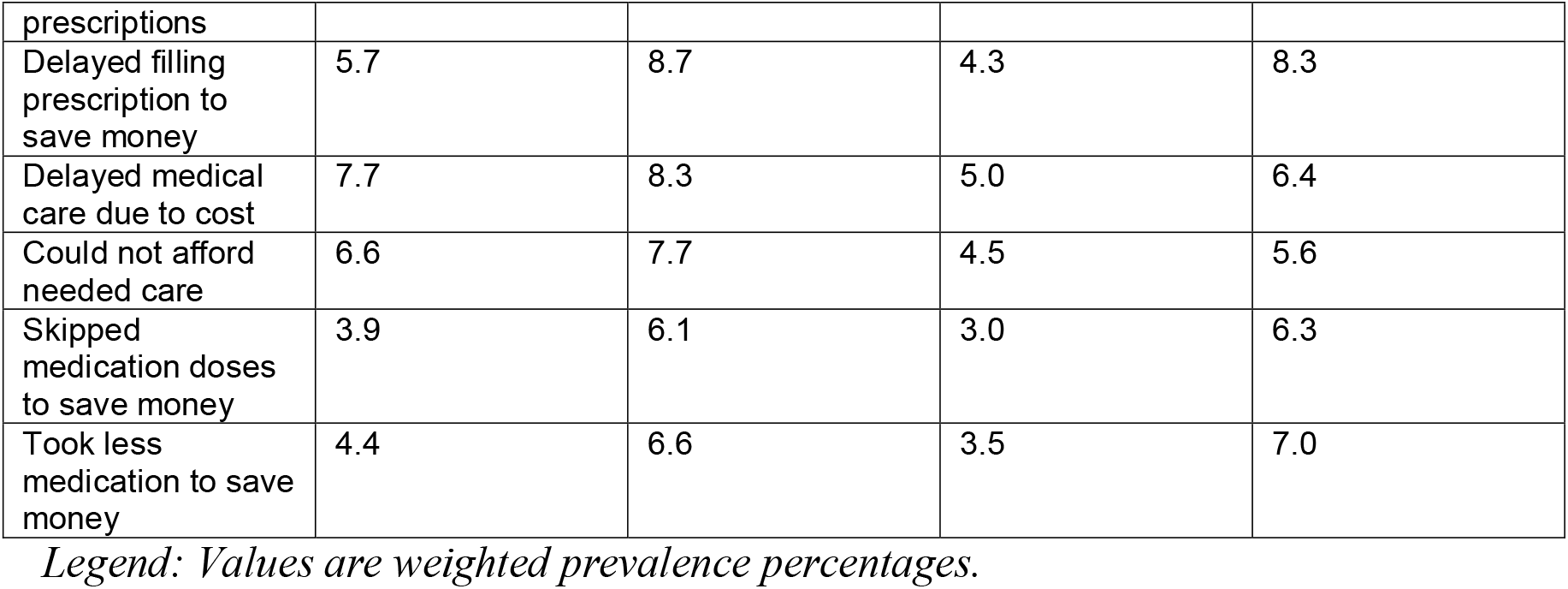
Weighted prevalence of financial-toxicity outcomes by phenotype group.

**Figure 1.**
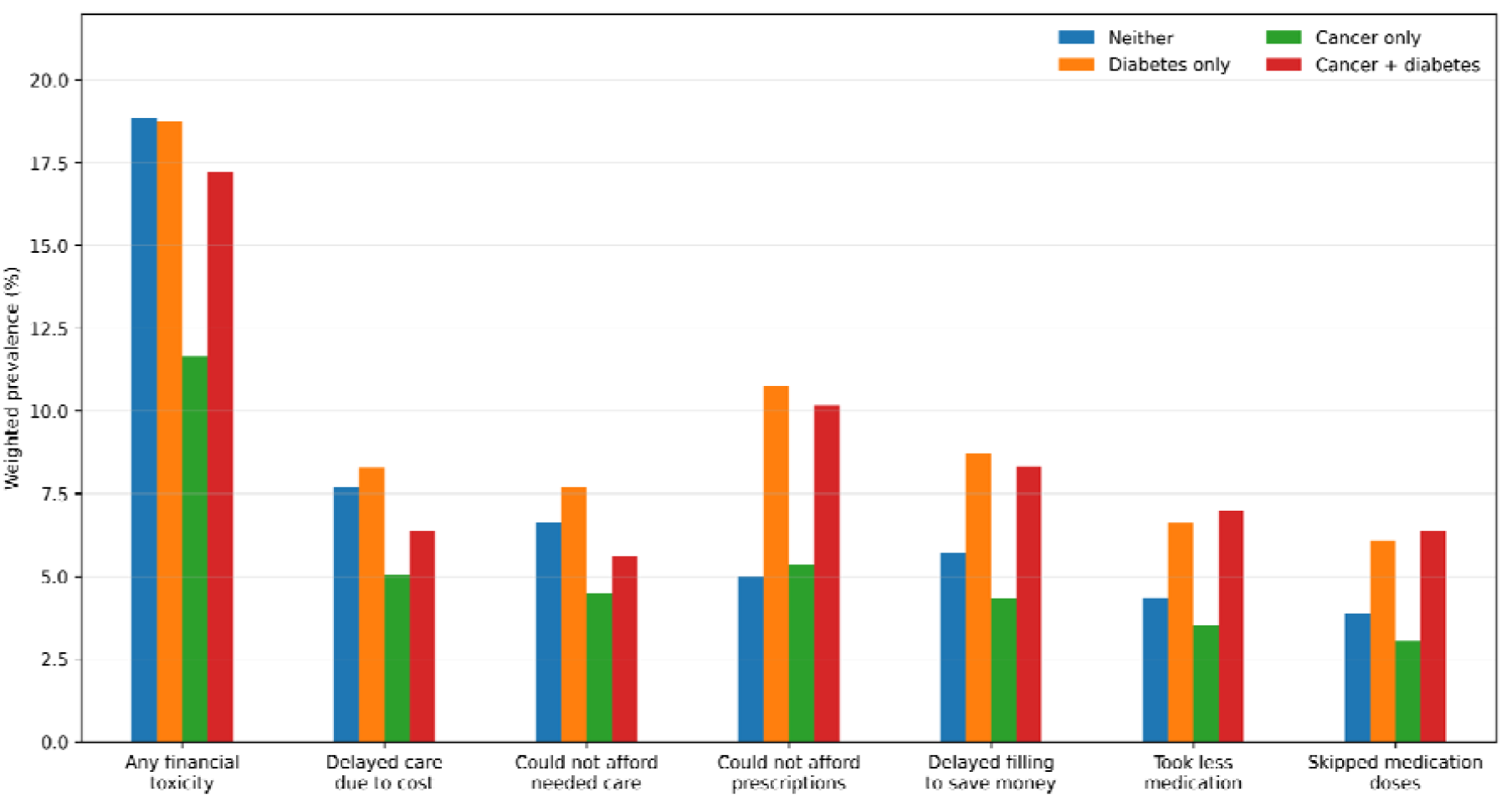
Weighted prevalence of financial-toxicity outcomes by cancer-diabetes phenotype.

Adults with both cancer and diabetes consistently showed high levels of prescription-related cost strain. The prevalence of being unable to afford prescriptions was 10.2% in the cancer-plus-diabetes group, compared with 10.7% in the diabetes-only group, 5.4% in the cancer-only group, and 5.0% in the neither-condition group. Delaying prescription filling to save money occurred in 8.3% of adults with cancer and diabetes, compared with 8.7% in diabetes only and 4.3% in cancer only. Taking less medication to save money occurred in 7.0% of the combined phenotype group, compared with 6.6% in diabetes only and 3.5% in cancer only.

Mean financial-toxicity burden followed a similar pattern. The mean number of financial-toxicity items was 0.28 in the neither group, 0.47 in the diabetes-only group, 0.25 in the cancer-only group, and 0.43 in the cancer-plus-diabetes group. The prevalence of having ≥2 financial-toxicity items was 7.8% in the neither group, 12.0% in diabetes only, 6.4% in cancer only, and 11.3% in cancer plus diabetes.

### Healthcare access and utilization across phenotype groups

Despite high financial burden, adults with both cancer and diabetes did not appear disconnected from care. The prevalence of having no usual place for care was lowest in the cancer-plus-diabetes group (1.3%), compared with 2.8% in diabetes only, 2.8% in cancer only, and 12.0% in the neither group. At the same time, the combined phenotype had the highest acute care use. Any emergency department visit was reported by 35.9% of adults with both cancer and diabetes, compared with 28.7% in diabetes only, 24.5% in cancer only, and 18.1% in the neither group. The prevalence of ≥2 emergency department visits was likewise highest in the cancer-plus-diabetes group at 15.8%. Preventive healthcare contact was also greatest in this group, with 87.6% reporting a wellness visit within the past year. Detailed healthcare access and utilization estimates are shown in Supplementary Table S7.

### Adjusted association between phenotype group and financial-toxicity severity

In adjusted analyses, diabetes appeared to be the principal driver of financial-toxicity severity. Relative to adults with neither condition, the adjusted prevalence ratio for having ≥2 financial-toxicity items was 1.80 (95% CI 1.66–1.96) for diabetes only and 1.90 (95% CI 1.63–2.21) for cancer plus diabetes, whereas cancer only was not significantly associated with greater severity (aPR 1.07, 95% CI 0.96–1.20). The distribution of FT severity across phenotype groups is shown in Supplementary Figure S1. In a sensitivity model additionally adjusted for food security and urban-rural classification, the combined cancer-plus-diabetes phenotype remained significantly associated with high FT severity (aPR 1.69, 95% CI 1.47– 1.95), whereas the cancer-only phenotype remained non-significant (aPR 1.09, 95% CI 0.98–1.21). Detailed estimates for the additional-adjustment model and the urban-rural stratified analysis are shown in Supplementary Tables S3 and S4.

### Cancer survivors: impact of diabetes on financial toxicity

Within cancer survivors, the crude prevalence of any financial toxicity was 11.6% in those without diabetes and 17.2% in those with diabetes. Similar differences were observed across all prescription-related outcomes. For example, the prevalence of being unable to afford prescriptions was 5.4% in cancer survivors without diabetes and 10.2% in those with diabetes, while delaying prescription filling to save money occurred in 4.3% and 8.3%, respectively.

In multivariable models adjusted for age, sex, race/ethnicity, education, poverty, insurance, region, BMI category, smoking, and survey year, diabetes among cancer survivors was associated with substantially greater cost-related burden. Compared with cancer survivors without diabetes, those with diabetes had higher prevalence of any financial toxicity (aPR 1.51, 95% CI 1.33–1.73), delayed medical care due to cost (aPR 1.47, 95% CI 1.14–1.88), inability to afford needed care (aPR 1.37, 95% CI 1.08–1.74), inability to afford prescriptions (aPR 1.71, 95% CI 1.40–2.08), delayed filling prescriptions to save money (aPR 1.77, 95% CI 1.42–2.21), taking less medication to save money (aPR 1.81, 95% CI 1.42–2.32), and skipping medication doses to save money (aPR 1.91, 95% CI 1.48–2.46) (Table 3; Figure 2).

**Table 3.** Adjusted prevalence ratios for financial-toxicity and healthcare-vulnerability outcomes among cancer survivors with versus without diabetes.

| Outcome | aPR (95% CI) | P value |
| --- | --- | --- |
| Any financial toxicity | 1.51 (1.33–1.73) | <0.001 |
| Delayed care due to cost | 1.47 (1.14–1.88) | 0.003 |
| Could not afford needed care | 1.37 (1.08–1.74) | 0.011 |
| Could not afford prescriptions | 1.71 (1.40–2.08) | <0.001 |
| Delayed filling prescription | 1.77 (1.42–2.21) | <0.001 |
| Took less medication | 1.81 (1.42–2.32) | <0.001 |
| Skipped medication doses | 1.91 (1.48–2.46) | <0.001 |
| No usual place for care | 0.35 (0.18–0.68) | 0.002 |
| Any ER visit | 1.35 (1.24–1.47) | <0.001 |
| ≥2 ER visits | 1.55 (1.32–1.83) | <0.001 |
| Wellness visit within past year | 1.13 (1.06–1.19) | <0.001 |
| Most recent visit was wellness | 0.99 (0.96–1.02) | 0.492 |

*Legend: Reference group: cancer survivors without diabetes. Models adjusted for age, sex, race/ethnicity, education, poverty, insurance, region, BMI category, smoking, and survey year. aPR, adjusted prevalence ratio; CI, confidence interval.*

**Figure 2.**
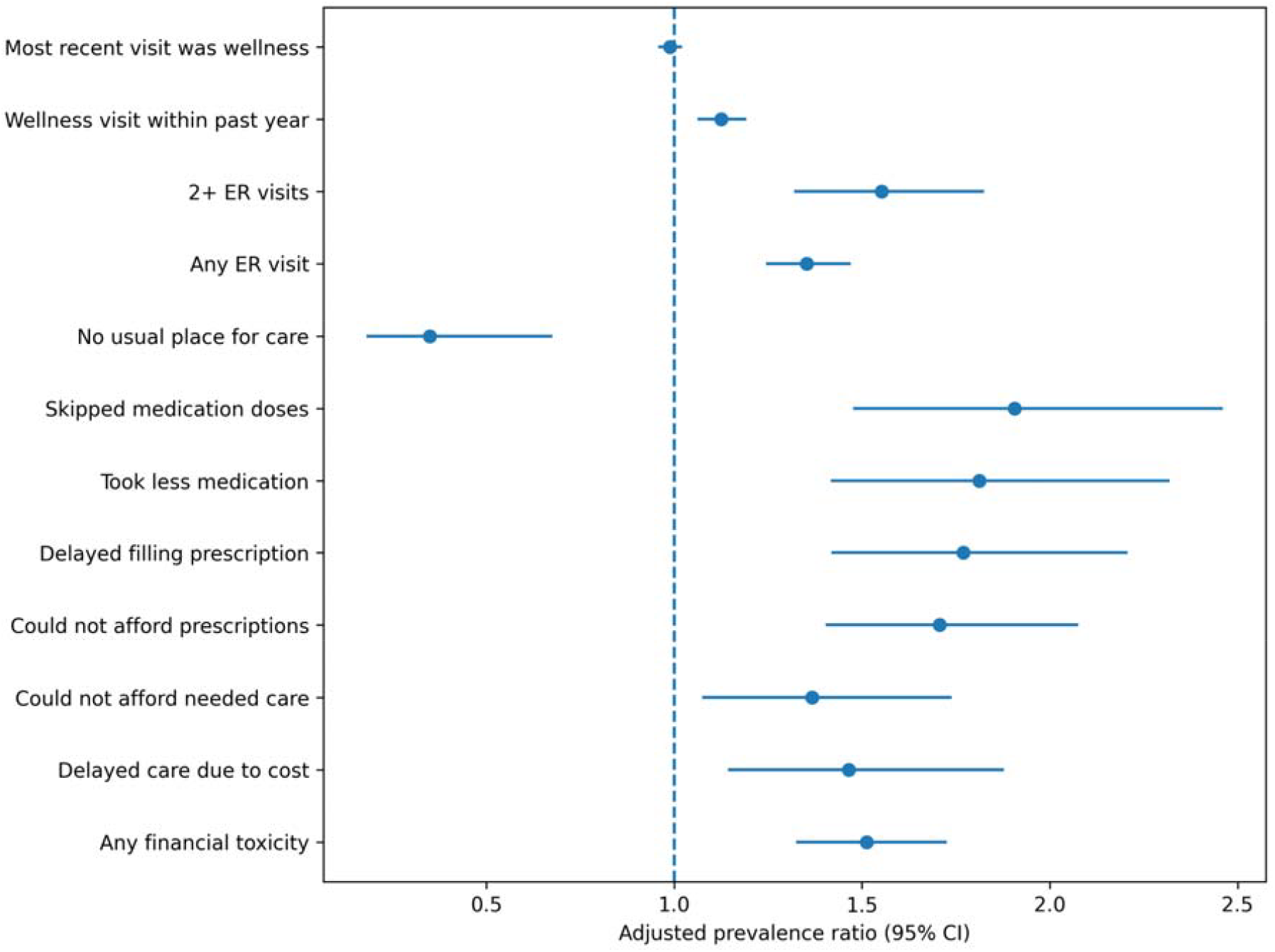
Adjusted prevalence ratios for key financial-toxicity and healthcare-vulnerability outcomes among cancer survivors with versus without diabetes.

### Cancer survivors: diabetes and healthcare use

Within cancer survivors, diabetes was also associated with greater healthcare utilization. Compared with cancer survivors without diabetes, those with diabetes had higher adjusted prevalence of any emergency department visit (aPR 1.35, 95% CI 1.24–1.47) and ≥2 emergency department visits (aPR 1.55, 95% CI 1.32–1.83). They were also more likely to report a wellness visit within the past year (aPR 1.13, 95% CI 1.06–1.19). In contrast, diabetes was associated with a lower prevalence of having no usual place for care (aPR 0.35, 95% CI 0.18–0.68). The prevalence of having the most recent doctor visit be a wellness visit did not differ materially by diabetes status (aPR 0.99, 95% CI 0.96–1.02).

Taken together, these findings suggest that cancer survivors with diabetes have frequent contact with the healthcare system, yet remain substantially more vulnerable to cost-related care disruption and prescription underuse.

### Food insecurity

Food insecurity also differed across phenotype groups. The prevalence of food insecurity was 7.5% in the neither group, 12.1% in diabetes only, 5.0% in cancer only, and 10.2% in cancer plus diabetes, indicating that the combined phenotype carried substantially greater social vulnerability than cancer alone (Supplementary Table S2).

### Subgroup analyses among cancer survivors

The association between diabetes and any financial toxicity among cancer survivors was consistent across nearly all subgroups examined. The association was present in both younger and older adults, but was stronger among those aged 65 years or older (aPR 1.80, 95% CI 1.48–2.19) than among those younger than 65 years (aPR 1.39, 95% CI 1.17–1.66). By sex, diabetes was associated with higher financial toxicity in both men (aPR 1.71, 95% CI 1.35–2.16) and women (aPR 1.49, 95% CI 1.28–1.74). By insurance status, the association remained significant among those with private insurance (aPR 1.44, 95% CI 1.18–1.76) and Medicare (aPR 1.76, 95% CI 1.41–2.19). Point estimates were also elevated in Medicaid and other/uninsured strata, although confidence intervals crossed the null. By poverty level, diabetes remained associated with higher financial toxicity across all income strata, including <100% FPL (aPR 1.52, 95% CI 1.10–2.11), 100%–199% FPL (aPR 1.52, 95% CI 1.24–1.86), and ≥200% FPL (aPR 1.59, 95% CI 1.30–1.93).

### Diabetes treatment intensity among cancer survivors

When cancer survivors with diabetes were stratified by current treatment status, a gradient of vulnerability emerged. Relative to cancer survivors without diabetes, the adjusted prevalence of any financial toxicity was higher in those with diabetes but no current medications reported (aPR 1.47, 95% CI 1.17–1.84), in those with non-insulin-treated diabetes (aPR 1.50, 95% CI 1.24–1.81), and in those with insulin-treated diabetes (aPR 1.56, 95% CI 1.26–1.92).

A similar pattern was observed for prescription affordability. The adjusted prevalence of being unable to afford prescriptions was 1.77-fold higher among those with diabetes and no current diabetes medications reported, 1.59-fold higher among those with non-insulin-treated diabetes, and 1.83-fold higher among those with insulin-treated diabetes, compared with cancer survivors without diabetes. For delayed care due to cost, the association was significant in non-insulin-treated diabetes (aPR 1.46, 95% CI 1.03–2.06) and insulin-treated diabetes (aPR 1.58, 95% CI 1.08–2.30), while the point estimate for those with no current diabetes medications reported was elevated but not statistically significant. For inability to afford needed care, the strongest association was again observed in insulin-treated diabetes, although this estimate was of borderline statistical significance (Figure 3; Supplementary Table S8).

**Figure 3.**
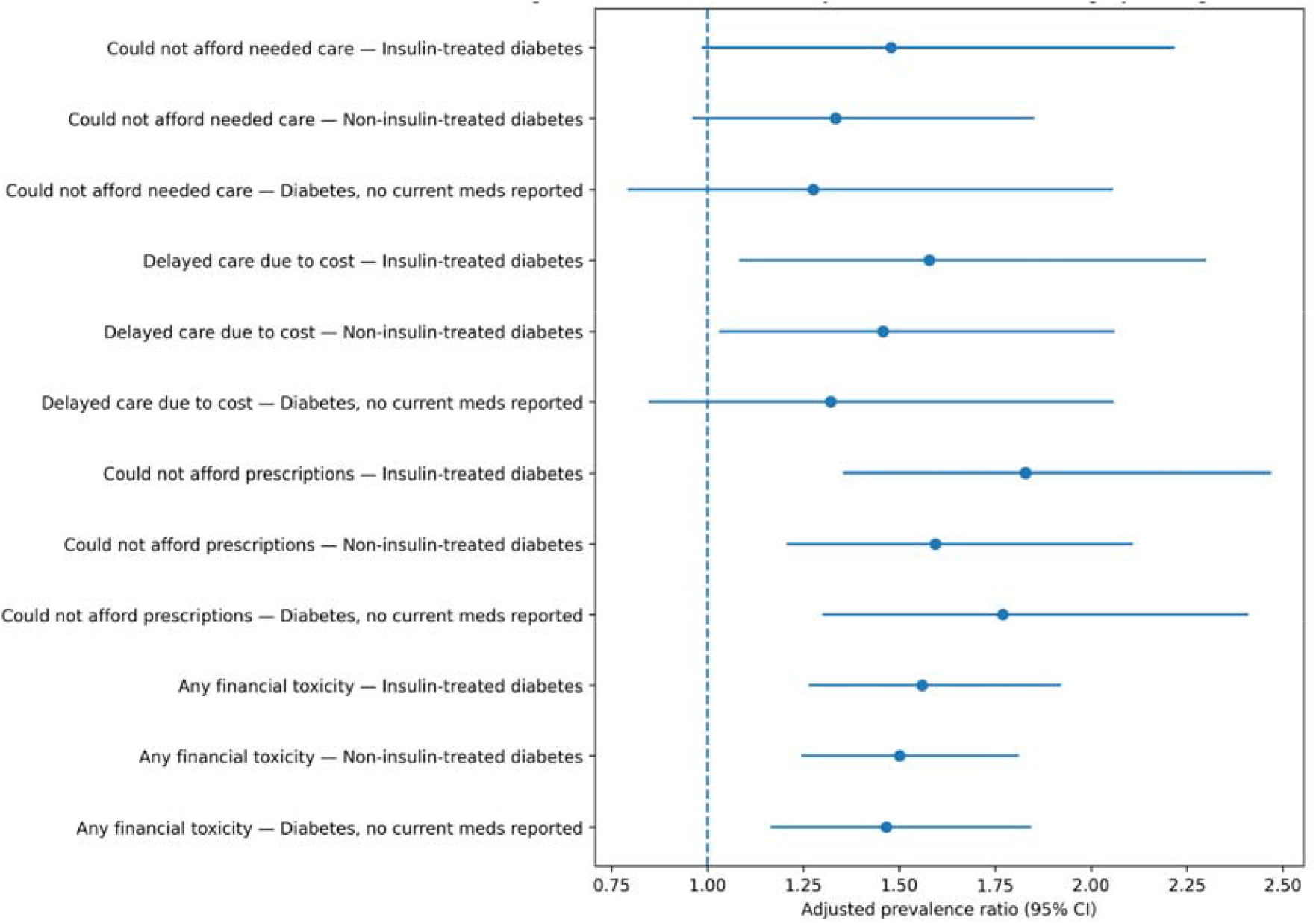
Adjusted prevalence ratios for selected financial-toxicity outcomes by diabetes treatment category among cancer survivors.

Overall, these findings suggest that financial vulnerability among cancer survivors is not only associated with diabetes itself, but may be greatest among those with more treatment-intensive disease. Exploratory site-specific analyses showed a similar direction of association across several major cancer types and are provided in Supplementary Table S5.

## Discussion

In this nationally representative study, cancer survivors with diabetes had substantially greater financial toxicity than cancer survivors without diabetes. This excess burden was seen not only for the composite outcome, but also across all prescription-related measures. These findings are consistent with the broader literature showing that financial hardship in survivorship commonly manifests as delayed or forgone care and medication nonadherence (1,2,15–20).

Our results also align with prior evidence that comorbidity worsens the economic burden of cancer survivorship. Rim et al reported in a systematic review that chronic conditions increase the economic burden of survivorship, while Fastiggi et al found that comorbidities were associated with greater financial hardship among adult cancer survivors (8,9). National studies by Zheng et al and Han et al further showed that medical financial hardship and financial sacrifice are common among survivors in the United States, and Yabroff et al demonstrated that financial hardship is associated with worse long-term outcomes, including mortality (16–18). The present study extends that literature by showing that diabetes specifically is associated with higher cost-related care disruption and prescription underuse even after adjustment for socioeconomic, insurance, and health factors.

Importantly, the combined phenotype remained significantly associated with high FT severity even after additional adjustment for food security and urban-rural classification. This suggests that the excess burden among cancer survivors with diabetes is not explained solely by broader social vulnerability or geography, but reflects a more specific pattern of compounded healthcare and medication burden.

One notable finding was the lower crude prevalence of FT in the cancer-only group relative to the neither-condition group. This likely reflects differences in age and insurance structure rather than lower true vulnerability, as the cancer-only group was older and more likely to have Medicare coverage. After adjustment, the apparent crude advantage of cancer-only status attenuated. In contrast, diabetes remained associated with higher FT, including among older adults and Medicare-insured survivors, suggesting that the cumulative out-of-pocket burden of diabetes management may persist despite insurance coverage.

The higher prevalence of food insecurity in the combined phenotype further supports the interpretation that cancer survivors with diabetes face layered material hardship beyond clinical multimorbidity alone.

The healthcare-contact findings are particularly important. Cancer survivors with diabetes were more likely to report wellness visits and emergency department use, yet they still had markedly higher financial toxicity. This suggests that the problem is not simply lack of healthcare contact. Rather, these survivors appear to remain engaged in care while still facing affordability barriers that disrupt recommended treatment and medication use. This interpretation is consistent with prior work describing the complexity of cancer-diabetes co-management. Pinheiro et al reported substantial patient-perceived challenges in managing both conditions during cancer treatment, while earlier work by Hershey et al and the systematic review by Vissers et al showed that cancer treatment can disrupt diabetes self-management and worsen patient-reported outcomes (5–7). Similarly, Jo et al reported greater healthcare use among adults with cancer and diabetes, and McDaniel et al found greater economic burden in Medicare beneficiaries with both conditions (10,11).

Our treatment-stratified results support the idea that burden is greatest when diabetes is more treatment-intensive. Insulin-treated survivors had the highest prevalence of financial toxicity and the strongest associations for prescription affordability and delayed care. This is clinically plausible, because treatment-intensive diabetes generally requires more medications, supplies, monitoring, and outpatient management. It is also consistent with the diabetes-oncology literature showing that cancer can adversely affect diabetes care quality and medication adherence. Griffiths et al found in a systematic review that cancer is associated with poorer adherence to diabetes medications, and Zanders et al reported a decline in adherence to glucose-lowering therapy after cancer diagnosis (21,22).

Another implication of our findings is that standard insurance coverage may not be sufficient to protect high-risk survivors from cost-related disruption. Although financial toxicity is often framed primarily as a problem of the uninsured, our subgroup analyses showed persistent excess burden among privately insured and Medicare-insured cancer survivors with diabetes. This observation is in line with prior survivorship research emphasizing that financial toxicity is shaped not only by insurance status, but also by treatment burden, work disruption, and accumulated out-of-pocket costs (1,2,16,18,23).

This study has limitations. NHIS is cross-sectional, so temporal directionality cannot be established. Cancer and diabetes were self-reported, and the public-use NHIS files do not fully characterize cancer type, stage, treatment, or time since diagnosis (12,14,24). We also measured financial toxicity using cost-related care disruption and medication underuse rather than direct out-of-pocket spending or debt. However, these are clinically meaningful indicators of affordability-related vulnerability and are well suited to population-based survey research (19,20,24,25).

Despite these limitations, this study has important strengths. It uses recent, nationally representative U.S. data, captures both general medical-care disruption and prescription-specific financial burden, and evaluates the coexistence of cancer and diabetes within a unified phenotype framework. Overall, the findings suggest that diabetes is not merely another comorbidity in survivorship, but a marker of sustained financial vulnerability.

## Conclusion

Cancer survivors with diabetes represent a high-risk financial-toxicity phenotype in the United States. Compared with cancer survivors without diabetes, they experience substantially greater cost-related care disruption and prescription underuse despite frequent healthcare contact, with the highest burden observed among insulin-treated survivors. Integrated survivorship and diabetes-care strategies may help reduce financial strain in this vulnerable population.

## Data Availability

All data produced in the present study are available upon reasonable request to the authors

## Author Contributions

J.T.: conceptualization, methodology, formal analysis, data curation, and writing—original draft. V.S., H.N., A.T., V.T., and A.S.: methodology, interpretation of data, and critical revision of the manuscript. All authors reviewed and approved the final manuscript.

## Data Availability

The data underlying this study are publicly available from IPUMS NHIS and the National Health Interview Survey documentation. Access to harmonized IPUMS NHIS files may require registration with IPUMS.

## Code Availability

Analytic code used for data cleaning and statistical analysis is available from the authors on reasonable request.

## Acknowledgments

None.

## References

1. Altice CK, Banegas MP, Tucker-Seeley RD, Yabroff KR. Financial Hardships Experienced by Cancer Survivors: A Systematic Review. JNCI J Natl Cancer Inst. 2016;109(2):djw205. doi:10.1093/jnci/djw205

2. Gordon LG, Merollini KMD, Lowe A, Chan RJ. A Systematic Review of Financial Toxicity Among Cancer Survivors: We Can’t Pay the Co-Pay. The Patient. 2017;10(3):295–309. doi:10.1007/s40271-016-0204-x

3. American Diabetes Association Professional Practice Committee. 1. Improving care and promoting health in populations: Standards of Care in Diabetes—2024. Diabetes Care. 2024;47(Suppl 1):S11–S19. doi:10.2337/dc24-S001

4. Khera R, Valero-Elizondo J, Das SR, et al. Cost-Related Medication Nonadherence in Adults With Atherosclerotic Cardiovascular Disease in the United States, 2013 to 2017. Circulation. 2019;140(25):2067–2075. doi:10.1161/CIRCULATIONAHA.119.041974

5. Pinheiro LC, Cho J, Rothman J, et al. Diabetes and cancer co-management: patient-reported challenges, needs, and priorities. Support Care Cancer. 2023;31(2):145. doi:10.1007/s00520-023-07604-x

6. Hershey DS, Tipton J, Given B, Davis E. Perceived Impact of Cancer Treatment on Diabetes Self-Management. Diabetes Educ. 2012;38(6):779–790. doi:10.1177/0145721712458835

7. Vissers PAJ, Falzon L, van de Poll-Franse LV, Pouwer F, Thong MSY. The impact of having both cancer and diabetes on patient-reported outcomes: a systematic review and directions for future research. J Cancer Surviv. 2016;10:406–415. doi:10.1007/s11764-015-0486-3

8. Rim SH, Guy GP, Yabroff KR, McGraw KA, Ekwueme DU. The impact of chronic conditions on the economic burden of cancer survivorship: a systematic review. Expert Rev Pharmacoecon Outcomes Res. 2016;16(5):579–589. doi:10.1080/14737167.2016.1239533

9. Fastiggi MJ, Sim J-A, Huang I-C. Association of co-morbidities with financial hardship in survivors of adult cancer. Support Care Cancer. 2021;29(12):7355–7364. doi:10.1007/s00520-021-06313-7

10. Jo A, Parikh S, Sawczuk N, Turner K, Hong YR. Health Care Use Among Cancer Patients With Diabetes, National Health and Nutrition Examination Survey, 2017–2020. Prev Chronic Dis. 2024;21:E58. doi:10.5888/pcd21.240066

11. McDaniel CC, Loh FE, Rockwell DM, McDonald CP, Chou C. Economic burden of diabetes among Medicare beneficiaries with cancer. J Pharm Health Serv Res. 2021;12(2):142–151. doi:10.1093/jphsr/rmab002

12. CDC. 2024 NHIS Questionnaires, Datasets, and Documentation. National Health Interview Survey. July 21, 2025. Accessed April 23, 2026. https://www.cdc.gov/nchs/nhis/documentation/2024-nhis.html

13. IPUMS NHIS. Accessed April 23, 2026. https://nhis.ipums.org/nhis/userNotes_weights.shtml

14. National Center for Health Statistics. 2024 National Health Interview Survey (NHIS) codebook for sample adult file. Centers for Disease Control and Prevention; 2025. Accessed March 26, 2026. https://ftp.cdc.gov/pub/Health_Statistics/NCHS/Dataset_Documentation/NHIS/2024/adult-codebook.pdf

15. Yabroff KR, Zhao J, Zheng Z, Rai A, Han X. Medical Financial Hardship among Cancer Survivors in the United States: What Do We Know? What Do We Need to Know? Cancer Epidemiol Biomarkers Prev. 2018;27(12):1389–1397. doi:10.1158/1055-9965.EPI-18-0617

16. Zheng Z, Jemal A, Han X, et al. Medical financial hardship among cancer survivors in the United States. Cancer. 2019;125(10):1737–1747. doi:10.1002/cncr.31913

17. Han X, Zhao J, Zheng Z, de Moor JS, Virgo KS, Yabroff KR. Medical Financial Hardship Intensity and Financial Sacrifice Associated with Cancer in the United States. Cancer Epidemiol Biomarkers Prev. 2020;29(2):308–317. doi:10.1158/1055-9965.EPI-19-0460

18. Yabroff KR, Han X, Song W, et al. Association of Medical Financial Hardship and Mortality Among Cancer Survivors in the United States. JNCI J Natl Cancer Inst. 2022;114(6):863–870. doi:10.1093/jnci/djac044

19. Thomy LB, Crichton M, Jones L, et al. Measures of financial toxicity in cancer survivors: a systematic review. Support Care Cancer. 2024;32(7):403. doi:10.1007/s00520-024-08601-4

20. Witte J, Mehlis K, Surmann B, et al. Methods for measuring financial toxicity after cancer diagnosis and treatment: a systematic review and its implications. Ann Oncol. 2019;30(7):1061–1070. doi:10.1093/annonc/mdz140

21. Griffiths RI, Keating NL, Bankhead CR. Quality of diabetes care in cancer: a systematic review. Int J Qual Health Care. 2019;31(2):75–88. doi:10.1093/intqhc/mzy124

22. Zanders MMJ, Haak HR, van Herk-Sukel MPP, van de Poll-Franse LV, Johnson JA. Impact of cancer on adherence to glucose-lowering drug treatment in individuals with diabetes. Diabetologia. 2015;58(5):951–960. doi:10.1007/s00125-015-3497-8

23. Mols F, Tomalin B, Pearce A, Kaambwa B, Koczwara B. Financial toxicity and employment status in cancer survivors: a systematic literature review. Support Care Cancer. 2020;28(12):5693–5708. doi:10.1007/s00520-020-05719-z

24. National Center for Health Statistics. 2024 National Health Interview Survey: Summary Health Statistics for the Sample Adult Population. Centers for Disease Control and Prevention; 2025. Accessed March 28, 2026. https://ftp.cdc.gov/pub/Health_Statistics/NCHS/Dataset_Documentation/NHIS/2024/adult-summary.pdf

25. Cohen RA, Mykyta L. Prescription Medication Use, Coverage, and Nonadherence Among Adults Age 65 and Older: United States, 2021-2022. Natl Health Stat Rep. 2024;(209):1–16. doi:10.15620/cdc/160016

