## Supplementary Files for "Diabetes as a Driver of Financial Toxicity Among U.S. Cancer Survivors: A Nationally Representative Analysis of NHIS 2021–2024"

This supplementary appendix contains additional tables and figures referenced in the main manuscript.

**Supplementary Table S1. Variable definitions for financial-toxicity outcomes**

| **Variable** | **Definition** | **Scale** |
| --- | --- | --- |
| Delayed medical care due to cost | DELAYCOST; delayed medical care due to cost in past 12 months | Binary (Yes/No) |
| Unable to afford needed medical care | YBARCARE; needed care but could not afford it | Binary (Yes/No) |
| Unable to afford prescription medicine | YBARMEDS; needed prescription but could not afford it | Binary (Yes/No) |
| Delayed filling prescription to save money | YDELAYMEDYR; delayed filling prescription | Binary (Yes/No) |
| Took less medication to save money | YSKIMPMEDYR; took less medication | Binary (Yes/No) |
| Skipped medication doses to save money | YSKIPMEDYR; skipped medication doses | Binary (Yes/No) |
| FT count | Summative number of affirmative FT items | Continuous 0-6 |
| FT severity | 0 items, 1 item, or 2+ items | Ordinal |

*Definitions of the six component items and derived financial-toxicity measures.*

**Supplementary Table S2. Food insecurity by cancer-diabetes phenotype**

| **Food security status** | **Cancer + diabetes** | **Cancer only** | **Diabetes only** | **Neither** |
| --- | --- | --- | --- | --- |
| Food insecure | 10 | 5 | 12 | 7 |
| Food secure | 90 | 95 | 88 | 93 |

*Values are weighted percentages.*

**Supplementary Table S3. FT severity model additionally adjusted for food security and urban-rural classification**

| **Phenotype vs neither** | **aPR (95% CI)** |
| --- | --- |
| Diabetes only | 1.61 (1.48–1.75) |
| Cancer only | 1.09 (0.98–1.21) |
| Cancer + diabetes | 1.69 (1.47–1.95) |

*Outcome shown is having ≥2 financial-toxicity items; reference group is neither cancer nor diabetes.*

**Supplementary Table S4. Urban-rural stratified FT severity model**

| **Setting** | **Phenotype vs neither** | **aPR (95% CI)** |
| --- | --- | --- |
| Metro | Diabetes only | 1.86 (1.68–2.04) |
| Metro | Cancer only | 1.11 (0.98–1.25) |
| Metro | Cancer + diabetes | 1.95 (1.64–2.31) |
| Nonmetro | Diabetes only | 1.44 (1.22–1.71) |
| Nonmetro | Cancer only | 1.17 (0.93–1.48) |
| Nonmetro | Cancer + diabetes | 1.78 (1.37–2.32) |

*Outcome shown is having ≥2 financial-toxicity items; reference group is neither cancer nor diabetes.*

**Supplementary Table S5. Exploratory financial-toxicity prevalence by major cancer type and diabetes status**

| **Cancer type** | **Diabetes, %** | **No diabetes, %** |
| --- | --- | --- |
| Bladder | 7.5 | 9.3 |
| Breast | 18.3 | 10.5 |
| Colorectal | 12.1 | 11.5 |
| Lung | 23.3 | 17.2 |
| Other/unspecified | 18.9 | 13.2 |
| Prostate | 12.7 | 4.7 |

*Values are weighted prevalence percentages.*

**Supplementary Table S6. Missingness for variables used in adjusted cancer-survivor models**

| **Variable** | **Missing n** | **Missing %** |
| --- | --- | --- |
| Any financial toxicity outcome | 1,321 | 8.7 |
| Diabetes status | 0 | 0.0 |
| Age | 0 | 0.0 |
| Sex | 0 | 0.0 |
| Race/ethnicity | 15 | 0.1 |
| Education group | 4,148 | 27.2 |
| Poverty group | 0 | 0.0 |
| Insurance group | 30 | 0.2 |
| US region | 0 | 0.0 |
| BMI category | 275 | 1.8 |
| Smoking status | 354 | 2.3 |
| Survey year | 0 | 0.0 |
| Food insecurity | 25 | 0.2 |
| Urban-rural classification | 0 | 0.0 |

*Cancer survivors, n=15,242. The fully adjusted main cancer-survivor model excluded 5,495 respondents because of missingness in one or more included variables, yielding a complete-case analytic sample of 9,747. The sensitivity model additionally including food insecurity and urban-rural classification excluded 5,508 respondents, yielding a complete-case analytic sample of 9,734.*

**Supplementary Table S7. Weighted prevalence of healthcare access and utilization outcomes by phenotype group**

| **Outcome** | **Cancer + diabetes** | **Cancer only** | **Diabetes only** | **Neither** |
| --- | --- | --- | --- | --- |
| 2+ ER visits in past year | 16 | 9 | 12 | 6 |
| Any ER visit in past year | 36 | 24 | 29 | 18 |
| Most recent doctor visit was wellness visit | 79 | 78 | 86 | 82 |
| No usual place for care | 1 | 3 | 3 | 12 |
| Wellness visit within past year | 88 | 74 | 80 | 47 |

*Values are weighted prevalence percentages. ER, emergency department.*

**Supplementary Table S8. Adjusted prevalence ratios by diabetes treatment category among cancer survivors**

| **Outcome** | **Diabetes treatment category** | **aPR (95% CI)** | **P value** |
| --- | --- | --- | --- |
| Any financial toxicity | Diabetes, no current meds reported | 1.47 (1.17–1.84) | 0.001 |
| Any financial toxicity | Non-insulin-treated diabetes | 1.50 (1.24–1.81) | <0.001 |
| Any financial toxicity | Insulin-treated diabetes | 1.56 (1.26–1.92) | <0.001 |
| Could not afford prescriptions | Diabetes, no current meds reported | 1.77 (1.30–2.41) | <0.001 |
| Could not afford prescriptions | Non-insulin-treated diabetes | 1.59 (1.21–2.11) | 0.001 |
| Could not afford prescriptions | Insulin-treated diabetes | 1.83 (1.35–2.47) | <0.001 |
| Delayed care due to cost | Diabetes, no current meds reported | 1.32 (0.85–2.06) | 0.219 |
| Delayed care due to cost | Non-insulin-treated diabetes | 1.46 (1.03–2.06) | 0.033 |
| Delayed care due to cost | Insulin-treated diabetes | 1.58 (1.08–2.30) | 0.018 |
| Could not afford needed care | Diabetes, no current meds reported | 1.28 (0.79–2.06) | 0.317 |
| Could not afford needed care | Non-insulin-treated diabetes | 1.33 (0.96–1.85) | 0.085 |
| Could not afford needed care | Insulin-treated diabetes | 1.48 (0.99–2.22) | 0.059 |

*Reference group: cancer survivors without diabetes. Models adjusted for age, sex, race/ethnicity, education, poverty, insurance, region, BMI category, smoking, and survey year.*

**Supplementary Figure S1. Distribution of financial-toxicity severity by phenotype group**

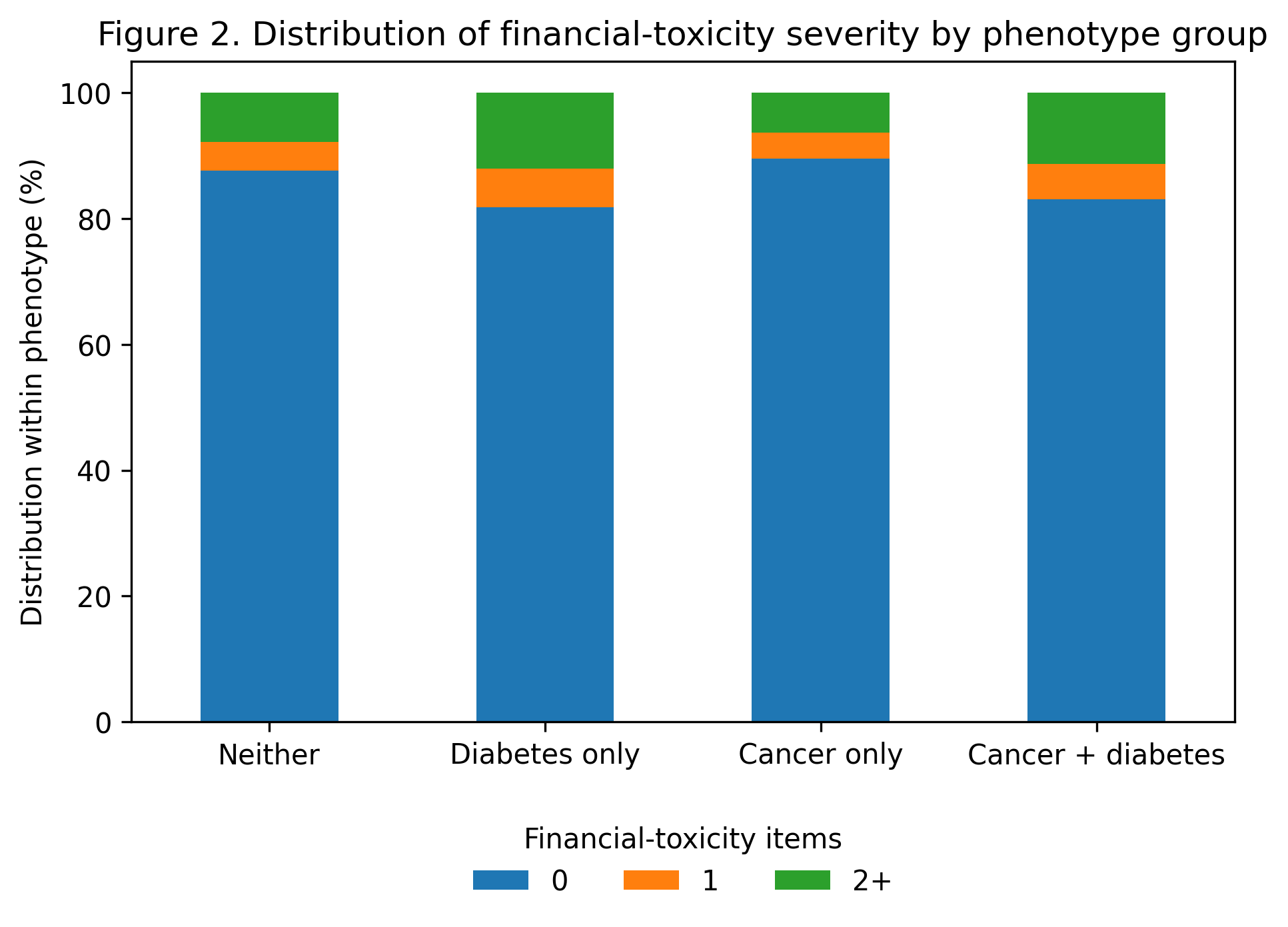

*Bars show the proportion with 0, 1, or ≥2 financial-toxicity items.*
